# Prevalence of SARS-CoV-2: An age-stratified, population-based, sero-epidemiological survey in Islamabad, Pakistan

**DOI:** 10.1101/2021.09.27.21264003

**Authors:** Mirza Amir Baig, Jamil Ahmed Ansari, Aamer Ikram, Mumtaz Ali Khan, Muhammad Salman, Zakir Hussain, Mirza Zeeshan Iqbal Baig, Ambreen Chaudhry, Muhammad Wasif Malik, Khurram Shahzad Akram, Asim Saeed, Muazzam Abbas Ranjha, Faisal Sultan, Sohail Sabir

## Abstract

**Introduction:** Serological surveys are valuable tools to evaluate the extent of disease transmission, measuring preventive effectiveness and proportion of asymptomatic individuals. This age-stratified, serological survey was aimed to measure the COVID-19 distribution and determinants in district Islamabad of Pakistan.

**Methodology:** Three-stage cluster sampling, using population proportionate to size technique, starting with a random number was used. A structured, pretested questionnaire was used after taking informed written consent, to gather demographic, risk factor information.

**Results:** Seroprevalence was found 16.5% (AR: 16.5%/100,000). The mean age was 35 (±16 Years). The majority were male (64%), self-employed (29%), and had primary level education (33%). The highest seroprevalence was found in the 21-30 years age group (24.8%) while the 41-50 years age group showed the highest attack rate (112.9/100,000 population). The proportion of the population tested that were asymptomatic was 69% (n=711) while the most frequently reported sign/symptom was cough (99%) followed by fever (20%). No known co-morbidity was reported in 86% (n=884) of respondents while hypertension remained the most reported condition (8%). High seroprevalence was observed in urban areas (12.3%) compared to rural union councils (6.4%). Visiting a house where COVID-19 case was isolated (OR 2, CI 1.38-2.84, *P*< 0.001), history of contact with a known case of COVID-19 (OR 1.42, CI 1.11-1.82, *P*=0.005), and attending a mass gathering (OR 1.21, CI: 1.02-1.42, p=0.02) were significant risk factors associated with contracting an infection. A Chi-Square test of independence showed significant protection while using regular hand hygiene practices (6.5; p<0.05) and regular usage of face masks (8.6; p<0.05).

**Conclusion:** Seroprevalence gives a direct estimation of population groups exposed to the virus. A remarkable difference in prevalence is found in urban and rural areas, extreme age groups, and socioeconomic statuses, suggesting targeted public health interventions. Sero-studies are affordable counterparts of molecular testing where quick estimation, prevention effectiveness, and data-driven public health policies are priorities.

## Introduction

In Pakistan, the first case of COVID-19 was identified on February 26, 2020 and by June 16, 2020, 148,921 cases and 2,839 deaths had been reported ^1^. Highest incidence was observed in Karachi, while Islamabad (capital of Pakistan) had a toll of 8,857 cases and 83 deaths during this time period ^2^. Confirmatory test for cases and their contacts (symptomatic or asymptomatic) is reverse transcriptase-polymerase chain reaction (real time RT-PCR), a gold standard confirmatory test. The detected number was largely influenced by the country’s daily testing capacity and active surveillance of contacts ^3^.

Epidemiological surveillance of confirmed COVID-19 cases, using RT-PCR, has provided a view of tip of the iceberg, referring to the spectrum of the disease from severe infection to asymptomatic transmission, and has been proven suitable for acute infections with signs and symptoms ^4^. Immune response to SARS-CoV-2 is varied and largely dependent on the severity of infection, host antibody response and is usually detectable after 7-14 days ^5 6^. Serological testing has advantage in disease surveillance in quantifying proportion of population exposed in a given period of time, where antibodies are used as a marker of partial or total immunity.

During this pandemic, seroprevalence studies has been carried out in confined populations, such as hospitals and nursing homes, health care workers or blood donors ^7^ where humoral immune response is measured but results are not generalizable. Sub national seroprevalence study results has been dissimilar in diverse areas, depending on magnitude of outbreak like; 1.5% of seropositivity was observed in Santa Clara, California while it was 32% in Boston which was considered to be a hot spot for COVID-19 outbreak ^8^, likewise, 7.1% in Atlanta USA, 9.6% in Wuhan, China and 21% in Guilan, Iran ^5^.

In Pakistan, attempts have been made to estimate the seroprevalence of COVID-19 in confined populations (police squads) ^9^ and in low and high transmission areas only ^10^ which are quite small and contained. A larger population based, age and gender stratified survey was needed to estimate the population at risk and factors associated with the exposures to SARS-CoV-2 ^11^. Since Islamabad Capital Territory (ICT) was the 4^th^ most affected city of Pakistan and to combat the current scenario, an enhanced surveillance strategy, based on population-based methods, was required ^12^. In an extensive literature review, use of serological testing of SARS-CoV-2 antibodies in general population, its role in estimation of burden of disease in different demographics during an epidemic, has been done but we could not find any evidence on the comparison of serology with molecular testing. Therefore, Field Epidemiology and Laboratory Training Program (FELTP) of Pakistan in collaboration with National Institute of Health (NIH) conducted a survey in district Islamabad, the capital of Pakistan, where age and gender stratification were used to help identify the exposed and at-risk population.

The primary objectives of the study were:

1. To estimate the extent of exposure to SARS-CoV-2 on demographic parameters
2. To estimate the proportion of asymptomatic infection in the population and prevalence of co-morbid conditions with SARS-CoV-2 exposure
3. To estimate the factors associated with SARS-CoV-2 exposure

## Methods

A community-based, age-stratified cross-sectional survey was conducted in district Islamabad in June 2020. Islamabad is divided into two administrative units; urban and rural. The World Health Organization’s (WHO) “*Population-based age-stratified sero-epidemiological investigation protocol for COVID-19 virus infection*” was followed to conduct this study ^13^.

Referring to 2017-18 census ^14^, total population of Islamabad is 200,657 with 336,182 households. The following formula was used to calculate the sample size:

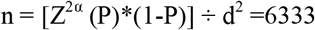

A three-staged stratified sampling procedure was adopted, using Population Proportionate to Size (PPS) at each stage. Union councils (UCs) are the lowest administrative units in a district. At the first stage, as per Pakistan Bureau of Statistics (PBS) ^14^ all urban and rural union councils (UCs) were enrolled (n=44) according to their population two each strata. The sample size was equally divided between rural (n=31, 49%), and urban (n=13, 51%) union councils for equal representation of both populations where urban UCs contain sectors and rural UCs contain villages in them. At the second stage, all the villages and sectors in these strata (rural and urban) were listed according to their population and again PPS sampling techniques was used. The sample was converted into households, and according to PBS, average five individuals live in a household. Finally, the number of households were enlisted, according to the data available at respective Town Management Office and Pakistan Bureau of Statistics, based on PPS techniques. A household was defined as *“a dwelling with a common kitchen or common opening onto a shared space*.*”* Using a systematic random sampling approach, a sampling interval was calculated by dividing the total number of households by number of households allocated per village/sector. All persons living in a household were included in the study for representation of all ages and genders (Figure 1).

**Figure 1:**
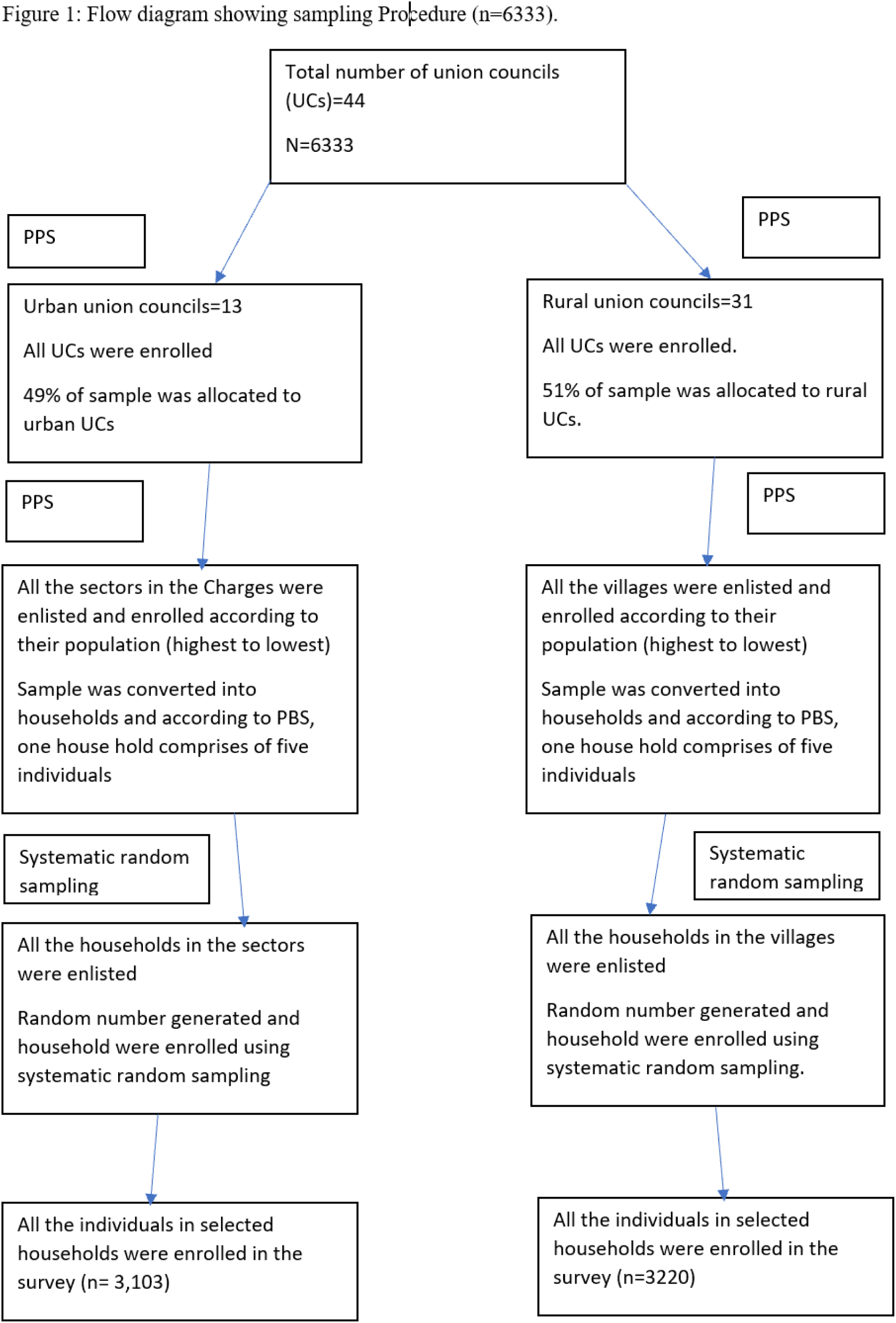
Flow diagram showing three-staged stratified sampling technique

A structured questionnaire was designed to collect demographics, disease status, clinical features, exposure information and other epidemiological parameters. Pilot testing was conducted for ensuring reliability and validity of the tool, in the neighboring district of Rawalpindi which guided further editing and improvement of the questionnaire.

Survey teams were trained on key components of the survey i.e., questionnaire administration, infection prevention and control (IPC) measures, sample collection, labelling, transportation and risk communication.

An institutional review board (IRB) approval was obtained from National Institute of Health (NIH). A consent form was developed and translated into local language for the purpose of enabling the respondents to understand their autonomous right to decide to participate in the study and announcing the beneficence and justice rendered in the study ^15^.

Strict infection control measures set forth by Public Health Laboratories Division, NIH were adopted by trained laboratory staff. Blood samples were collected from participants upon recruitment in the investigation for antibody testing with chemiluminescent immunoassay (CMI). CE/FDA approved in vitro diagnostic kits for ELISA were used (Abbot’s Diagnostic IgG/IgM RTD). Nasopharyngeal swabs were also collected from symptomatic and/or high-risk respondents (age>60 years or have contact with COVID-19 confirmed case).

Data was coded to mask identities of respondents, double entered, validated and password protected at central office. All hard copies were kept in the lock and key. Response rate was 100% and the missed entries on participants’ information were completed by follow-up calls.

Statistical analyses were done using Epi Info-7 software. Frequencies, rates and ratios were calculated for demographics, and prevalence was computed. Crude attack rates, age, gender and area-specific attack rates were determined. Bivariate and multivariate analyses were carried out for risk factors at 95% confidence interval (CI) and a p-value less than 0.05. Chi-Square test was used to evaluate the significance of protective variables.

## Results

A total of 6,333 individuals were approached at their doorsteps, 100% provided consent and blood samples. No test results were inconclusive. Of 6,333 individuals enrolled, 1033 (16.3%) showed a reactive antibody test. Mean age of all seropositive individuals was 35 years (±16 years). Majority of seropositive individuals were male (64%), had at least primary level education (88%) and were self-employed (59%) (Table 1).

**Table-1:** Summary statistics of seropositive individuals for SARS-CoV-2 in district Islamabad (June 2020; n=1033)

|  | Cases (n) | Frequency<br>(%) | 95% CI |
| --- | --- | --- | --- |
| Gender |  |  |  |
| Male | 665 | 64 | 61-67 |
| Female | 368 | 36 | 33-38 |
| Employment status |  |  |  |
| Self employed | 605 | 59 | 55.5-61.5 |
| Unemployed | 294 | 29 | 25.8-31.3 |
| Government employed | 134 | 13 | 11-15.2 |
| Education |  |  |  |
| No formal education | 125 | 12 | 10-14 |
| Primary | 338 | 33 | 30-36 |
| Secondary | 15 | 1 | 0.9-2.4 |
| Under graduate | 295 | 29 | 25.9-31.4 |
| Graduate | 6 | 1 | 0.3-1.3 |
| Post graduate | 250 | 24 | 22-27 |

Highest seroprevalence was found in 21-30 years age group (24.8%) while highest attack rate was found in 41-50 years (11.3/10,000 population) (Table 2).

**Table-2:**
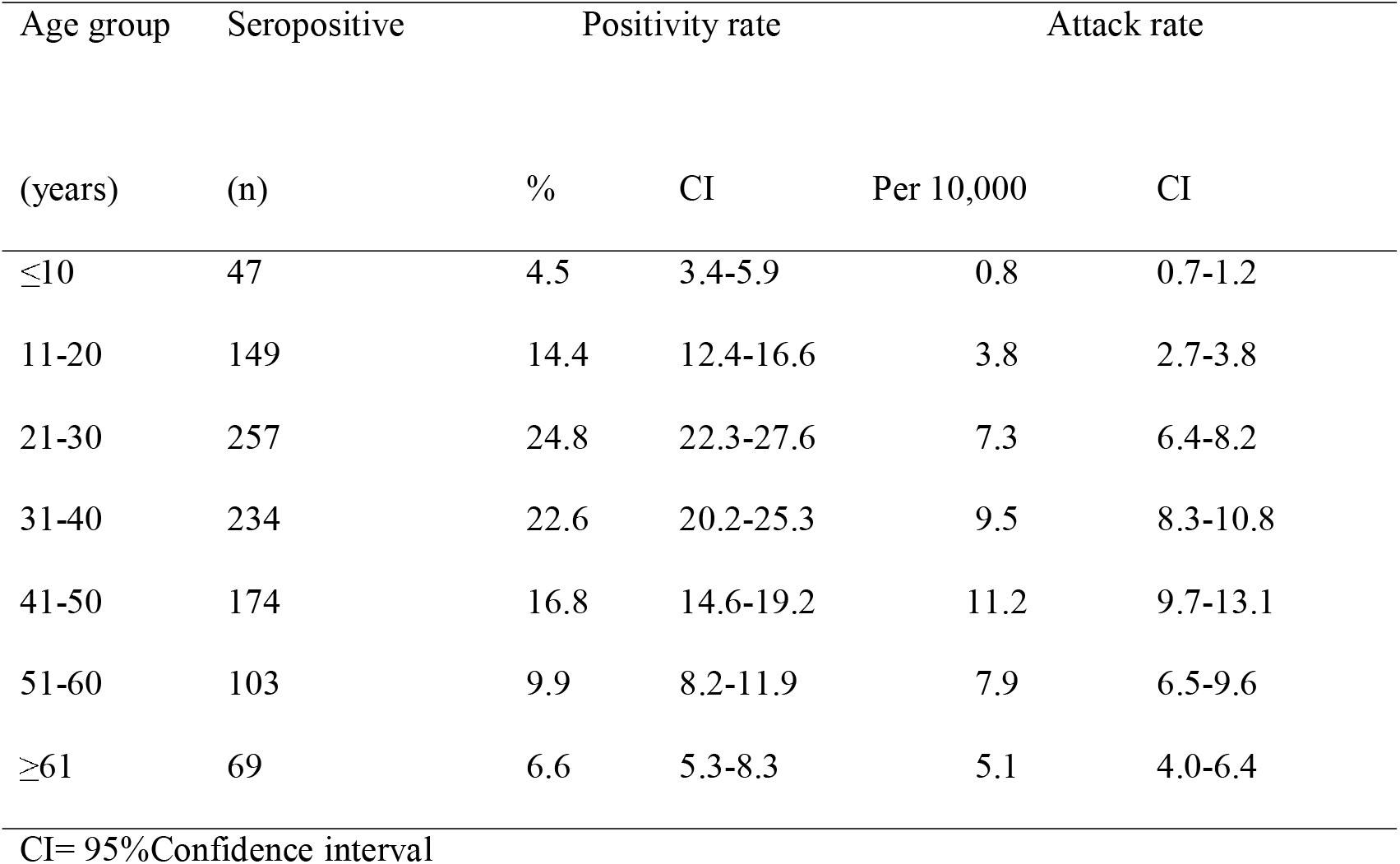
Age distribution of sero-positivity and attack rates for SARS-CoV-2 among residents of district Islamabad, June 2020 (n=1033)

Of all seropositive individuals, 711 (69%) were found asymptomatic within previous 14 days. Among symptomatic individuals, cough was the most reported symptom (99%) followed by fever (20%) and sore throat (12%). Among all respondents, 149 (14%) reported at least one known co-morbid condition and the most frequently reported co-morbidity was Hypertension (8%) followed by Diabetes Mellitus (7%) (Table 3).

**Table-3:** Clinical characteristics of sero-positive individuals for SARS-CoV-2 in district Islamabad, June 2020 (n=1033)

|  | Numbers<br>(n) | Frequency<br>(%) | 95% CI |
| --- | --- | --- | --- |
| Signs and symptoms ☐ |  |  |  |
| Cough | 319 | 99 | 97.7-99.2 |
| Fever | 202 | 20 | 17.2-22.1 |
| Sore throat | 119 | 12 | 9.7-13.6 |
| Headache | 109 | 11 | 8.8-12.5 |
| Shortness of breath | 67 | 7 | 5.1-8.1 |
| Diarrhea | 58 | 6 | 4.3-7.1 |
| Runny nose | 49 | 5 | 3.6-6.2 |
| Co-morbidities |  |  |  |
| No known co-morbidity | 884 | 86 | 83.3-87.5 |
| Hypertension | 79 | 8 | 6.2-9.43 |
| Diabetes Mellitus | 74 | 7 | 5.7-8.9 |
| IHD | 30 | 3 | 2.0-4.1 |
| CLD | 14 | 1 | 0.2-2.2 |
| CRD | 13 | 1 | 0.7-2.1 |
| Asthma | 10 | 1 | 0.5-1.8 |
| Chronic Lung Disease | 5 | 0.5 | 0.2-1.1 |

CI=confidence interval, IHD=Ischemic health disease, CLD=Chronic liver disease,
CRD=Chronic renal disease
☐ Self-reported signs and symptoms

Survey was conducted in 44 geographic localities of district Islamabad. High seroprevalence was found in urban union councils i.e., 12.3% in Charge13 (Sector: H-10 & I-10), 6.9% in charge 6 (Sector: F-6, 7 and Pir Sohawa) compared to lower seroprevalence in rural UCs (Alipur-6.4%, Tarlai Khurd-6.4%).

Out of 1033 seropositive respondents, 42/1033 (4 %) reported that they have visited a house where COVID-19 case was isolated (OR 2.0, CI: 1.38-2.84, p-0.001), 88/1033 (9%) had contact with a known case (OR 1.4, CI: 1.11-1.82, p-0.004), 216/1033 (21%) responded that they attended a mass gathering (OR 1.2, CI: 1.1-1.4, p-0.02) (Table 4).

**Table-4:** Association of risk factors with seroprevalence for SARS-CoV-2 among residents of district Islamabad, June 20202. (n=1033)

|  | Seropositive<br>(N) | Seronegative<br>(N) | OR | 95% CI | P-value |
| --- | --- | --- | --- | --- | --- |
| Visited a house where COVID-19 case was isolated |  |  |  |  |  |
| Yes | 42 | 111 | 2.0 | 1.38-2.84 | 0.001 |
| No | 991 | 5189 |  |  |  |
| Hospitalization |  |  |  |  |  |
| Yes | 4 | 12 | 1.7 | 0.50-5.30 | 0.4 |
| No | 1029 | 5288 |  |  |  |
| Contact with a known case |  |  |  |  |  |
| Yes | 88 | 325 | 1.4 | 1.11-1.82 | 0.02 |
| No | 945 | 4975 |  |  |  |
| Visited COVID-19 treatment facility |  |  |  |  |  |
| Yes | 27 | 110 | 1.3 | 0.82-1.93 | 0.27 |
| No | 1006 | 5190 |  |  |  |
| Attended a mass gathering |  |  |  |  |  |
| Yes | 216 | 952 | 1.2 | 1.11-1.41 | 0.34 |
| No | 817 | 4348 |  |  |  |
| Travelled abroad within 14 days |  |  |  |  |  |
| Yes | 32 | 196 | 0.8 | 0.52-1.23 | 0.34 |
| No | 1006 | 5190 |  |  |  |

OR=Odds ratio, CI=confidential interval,

Multi-variate analysis showed that visiting a house where COVID-19 case was isolated (aOR 2, CI: 1.38-2.84, p-value 0.001) was a significant risk factor associated with contracting an infection (Table 5).

**Table-5:** Multi-variate analysis of risk factors associated with seroprevalence for SARS-CoV-2 among residents of district Islamabad June 2020 (n=6333)

| Risk factors | Seropositive<br>(N) | Seronegative<br>(N) | aOR | 95% CI | p-value |
| --- | --- | --- | --- | --- | --- |
| Visited a house where<br>COVID-19 case was<br>isolated |  |  |  |  |  |
| Yes | 42 | 111 | 2.0 | 1.38-2.84 | 0.001 |
| No | 991 | 5189 |  |  |  |
| History of contact with<br>a known case |  |  |  |  |  |
| Yes | 88 | 325 | 1.4 | 1.11-1.82 | 0.005 |
| No | 945 | 4975 |  |  |  |
| Attended a mass<br>gathering |  |  |  |  |  |
| Yes | 216 | 952 | 1.2 | 1.02-1.42 | 0.02 |
| No | 817 | 4348 |  |  |  |

aOR=Adjusted odds ratio,

A Chi-Square test of independence indicated that the practices of regular use of a face mask and regular hand washing and/or hand sanitizing were correlated with lower seroprevalence (Table 6).

**Table-6:** Association of preventive measures and SARS-CoV-2 exposure among residents of district Islamabad, June 2020 (n=6333)

| Preventive measures | SARS-CoV-2<br>Positive (N) | SARS-CoV-2<br>Negative (N) | Chi-Square<br>Value | P-value |
| --- | --- | --- | --- | --- |
| Regular use of face mask |  |  |  |  |
| Regularly | 716 | 3423 | 8.6 | 0.03 |
| Occasionally | 176 | 1014 |  |  |
| No | 141 | 863 |  |  |
| Hand Washing/Hand Sanitizing |  |  |  |  |
| Regularly | 812 | 4292 | 6.5 | 0.01 |
| Occasionally | 131 | 661 |  |  |
| No | 90 | 347 |  |  |

Among all respondents 458/1033 (44%) were tested for RT-PCR and positivity rate was 14% (n=64).

## Discussion

Our results showed that the antibody response against COVID-19 was 16%, which translates to about 300,000 individuals being exposed to SARS-CoV-2 in district Islamabad. This should be constructed as this is not a reflection of cases and only depicts exposed individuals. It should be realized that these results were obtained through detecting antibodies against the SARS-CoV-2 virus which is not qualified as a diagnostic test for COVID-19. Confirmed cases can only be labelled through RT-PCR testing.

The results are in concordance with surveys conducted in Spain ^16^ where similar patterns has been documented in the general population and few other surveys conducted in Pakistan in localized and confined communities ^11^. Some small scale studies have also shown low seroprevalence with certain sampling biases such as enrolling only adult population ^17^. Our study displayed higher positivity rate among males and entrepreneurs which explains the outdoor activities related to these population groups even amidst series of smart lock downs in the district as well as skewed participation of males. On the other hand, low prevalence in extreme age groups has been observed (lower than 10 years and more than 60 years of age) which has been indicated in other studies also, conducted in developed countries including China, which are comparable with our findings (4.5 % and 6.6% respectively) ^18^.

We found 69% of seropositive individuals were asymptomatic, endorsing the outcome of various researches on stealth disease transmission ^19^ and a varied spectrum of signs and symptoms in the community and hospitalized individuals. This wide spectrum of clinical presentation has led to delayed diagnosis, delay in reaching health care facility and seeking medical care on one hand while undue fear, stigma and panic in society on the other hand ^20^. This high seroprevalence in asymptomatic individuals confirms the need for a greater serosurvey.

Likewise, our study has shown that more than half of responders did not declare a known co-morbid condition other than COVID-19, contrary to many studies done in hospitalized and older aged individuals who presented with a co-morbid condition with high fatality outcomes. Since we conducted this survey on general population so the results are difficult to compare with researches done in hospitalized and nursing home settings ^21^. We identified that among reported co-morbid conditions, hypertension and type-2 diabetes mellitus remained the most frequently reported one. A high prevalence of these non-communicable diseases in our communities has been documented with a maximum prevalence rate of 40% whereas some researchers suggest that more than half of them go undiagnosed due to low health services utility and health care access ^22^. Hence forth, coexistence of COVID-19 with aforementioned NCDs is not unusual. Rural population, though appears less affected, is comprised of a comparatively larger number of elderly people who are disproportionately impacted by the disease. The most germane space has been identified in urban areas but the rural areas of the district Islamabad seem more vulnerable in relative to age distribution.

Our results afford a strong evidence for smart lock down success instigated by government of Pakistan ^23^ where a significant association of exposure to COVID-19 with visiting a house where a COVID-19 case has been isolated, attending a mass gathering and having an epidemiological link, has been discerned. Equally, the strong association with the much advertised and advocated preventive measures has also been established ^24^. Therefore, this population based, age stratified, sero-epidemiological study has provided a robust evidence on the proportion of Islamabad (ICT) population exposed.

As RT-PCR is gold standard for the detection of disease, sero-testing can only complement the much resource limited and patchy surveillance of the low and middle income countries like Pakistan in an outbreak settings ^25^. Here, it is pertinent to mention that, a serological test cannot replace the confirmatory test, neither the results of one district be generalized across the country. Based on population distribution in terms of age, literacy rate and access to media and health care facilities, results of such sero-surveys may contrast. It has been recognized that the recommendations issued by WHO in March 2020 on “test, test, and test” ^26^ led to a huge burden on health system. Alternatively, rapid antigen/antibody detection tests were opted for many countries irrespective of their economic status, and are now recommended for quick and affordable point-of-care testing to triage exposed individuals. Antibody test detects the host response to virus which may take several days to develop, also it cannot distinguish between acute or previous infection ^27^. Some researchers have documented that high antibody response is associated with the severe disease. Low concurrent antigen and antibody detection, determined in our survey, suggests an exclusion of severely diseased from general population as they require hospitalization ^28^.

## Limitations

This survey, conducted in one district, shows seroprevalence of the same population and surveys using similar techniques in different populations may project different seroprevalence rates depending on the varied demographics, preventive measures taken, lock down policies and extent of exposure to SARS-CoV-2. Serological surveys give only the exposure status of population to SARS-CoV-2 and hence, should not be considered as their disease status. It is important to note that a greater number of males participated in the study compared to females.

## Conclusion

Diagnostics play a crucial role in epidemic response and so is the case with COVID-19. Rapid serology test is a scalable and affordable, population-based testing to estimate the extent of SAR-CoV-2 circulation in the community. Not only it gauges the population at risk, it fortifies the surveillance system to monitor trends over time. Based on our findings, rapid serological diagnostic testing is recommended in early and recession phase of outbreak to identify hot spots as well as measure the prevention effectiveness during high transmission phase. For astute distribution of limited public health resources, and to slacken off the burden on health care facilities, a rapid point-of-care testing will support in identifying target population and areas for smart lock down. Henceforth, implementation of testing tracking and quarantine (TTQ) strategy and achieving a data driven outbreak containment policies will be facilitated.

## Data Availability

The data will be available one request from the first author via email

## Financial support

Authors declare no financial support

## Conflict of interest

Authors declare no conflict of interest

## Ethical Approval

Institutional review Board of National Institute of Health Pakistan

## Notes

### Competing Interest Statement

The authors have declared no competing interest.

### Clinical Trial

NA

### Author Declarations

An institutional review board (IRB) approval was obtained from National Institute of Health Pakistan

## References

1. COVID-19 Health Advisory Platform by Ministry of National Health Services Regulations and Coordination. http://covid.gov.pk/. Accessed August 24, 2020.

2. Pakistan COVID-19 Corona Tracker. https://www.coronatracker.com/country/pakistan/. Accessed September 1, 2020.

3. Sethuraman N, Jeremiah SS, Ryo A. Interpreting Diagnostic Tests for SARS-CoV-2. JAMA - J Am Med Assoc. 2020;323(22):2249–2251. doi:10.1001/jama.2020.8259

4. Corman VM, Landt O, Kaiser M, et al. Detection of 2019 novel coronavirus (2019-nCoV) by real-time RT-PCR. Eurosurveillance. 2020. doi:10.2807/1560-7917.ES.2020.25.3.2000045

5. Lai AL, Millet JK, Daniel S, Freed JH, Whittaker GR. Testing for SARS-CoV-2 (COVID-19): a systematic review and clinical guide to molecular and seological in-vitro diagnostic assay. Lancet. 2020;395(April):1315.

6. Zhao J, Yuan Q, Wang H, et al. Antibody responses to SARS-CoV-2 in patients of novel coronavirus disease 2019. Clin Infect Dis. March 2020. doi:10.1093/cid/ciaa344

7. Valenti L, Bergna A, Pelusi S, et al. SARS-CoV-2 seroprevalence trends in healthy blood donors during the COVID-19 Milan outbreak. medRxiv. 2020:2020.05.11.20098442. doi:10.1101/2020.05.11.20098442

8. Madsen T, Levin N, Niehus K, et al. Prevalence of IgG antibodies to SARS-CoV-2 among emergency department employees. Am J Emerg Med. 2020. doi:10.1016/j.ajem.2020.04.076

9. Chughtai OR, Batool H, Khan MD, Chughtai AS. Frequency of COVID-19 IgG Antibodies among Special Police Squad Lahore, Pakistan. J Coll Physicians Surg Pak. 2020;30(7):735–739. doi:10.29271/jcpsp.2020.07.735

10. Nisar I, Ansari Msc N, Amin Msc M, et al. Serial population based serosurvey of antibodies to SARS-CoV-2 in a low and high transmission area of Karachi, Pakistan. medRxiv. August 2020:2020.07.28.20163451. doi:10.1101/2020.07.28.20163451

11. Javed W, Bin Baqar J, Hussain S, Abidi B, Farooq W. Sero-prevalence Findings from Metropoles in Pakistan: Implications for Assessing COVID-19 Prevalence and Case-fatality within a Dense, Urban Working Population. medRxiv. August 2020:2020.08.13.20173914. doi:10.1101/2020.08.13.20173914

12. World Health Organization. Covid 19 Strategy Update.; 2020.

13. World Health Organization. Population-based age-stratified seroepidemiological investigation protocol for COVID-19 virus infection. World Heal Organ. 2020.

14. Pakistan Bureau of Statistics. http://www.pbs.gov.pk/. Accessed August 26, 2020.

15. Belmont Report - an overview | ScienceDirect Topics. https://www.sciencedirect.com/topics/medicine-and-dentistry/belmont-report. Accessed August 25, 2020.

16. Pollán M, Pérez-Gómez B, Pastor-Barriuso R, et al. Prevalence of SARS-CoV-2 in Spain (ENE-COVID): a nationwide, population-based seroepidemiological study. Lancet. 2020;0(0). doi:10.1016/S0140-6736(20)31483-5

17. Sood N, Simon P, Ebner P, et al. Seroprevalence of SARS-CoV-2-Specific Antibodies among Adults in Los Angeles County, California, on April 10-11, 2020. JAMA - J Am Med Assoc. 2020. doi:10.1001/jama.2020.8279

18. Li W, Zhang B, Lu J, et al. The characteristics of household transmission of COVID-19. Clin Infect Dis. April 2020. doi:10.1093/cid/ciaa450

19. Huang C, Wang Y, Li X, et al. Clinical features of patients infected with 2019 novel coronavirus in Wuhan, China. Lancet. 2020;395(10223):497–506. doi:10.1016/S0140-6736(20)30183-5

20. Rothe C, Schunk M, Sothmann P, et al. Transmission of 2019-NCOV infection from an asymptomatic contact in Germany. N Engl J Med. 2020;382(10):970–971. doi:10.1056/NEJMc2001468

21. Moccia F, Gerbino A, Lionetti V, et al. COVID-19-associated cardiovascular morbidity in older adults: a position paper from the Italian Society of Cardiovascular Researches. GeroScience. 2020;42(4):1021–1049. doi:10.1007/s11357-020-00198-w

22. Shah N, Shah Q, Shah AJ. The burden and high prevalence of hypertension in Pakistani adolescents: A meta-analysis of the published studies. Arch Public Heal. 2018;76(1). doi:10.1186/s13690-018-0265-5

23. Pakistan among pioneers of “smart lockdown” approach, says PM Imran - Pakistan - DAWN.COM. https://www.dawn.com/news/1561766. Accessed August 25, 2020.

24. Coronavirus Disease 2019 (COVID-19) | CDC. https://www.cdc.gov/coronavirus/2019-ncov/index.html. Accessed August 24, 2020.

25. Nkengasong J. Let Africa into the market for COVID-19 diagnostics. Nature. 2020;580(7805):565–565. doi:10.1038/d41586-020-01265-0

26. WHO Director-General’s opening remarks at the media briefing on COVID-19 - 16 March 2020. https://www.who.int/dg/speeches/detail/who-director-general-s-opening-remarks-at-the-media-briefing-on-covid-19 16-march-2020. Accessed August 25, 2020.

27. He X, Lau EHY, Wu P, et al. Temporal dynamics in viral shedding and transmissibility of COVID-19. Nat Med. 2020;26(5):672–675. doi:10.1038/s41591-020-0869-5

28. Qu J, Wu C, Li X, et al. Profile of IgG and IgM antibodies against severe acute respiratory syndrome coronavirus 2 (SARS-CoV-2). Clin Infect Dis. April 2020. doi:10.1093/cid/ciaa489

